# Predication of Pandemic COVID-19 situation in Maharashtra, India

**DOI:** 10.1101/2020.04.10.20056697

**Authors:** Sunny Kumar

## Abstract

Presently, the world is infected by COVID 19 virus which has created an emergency for public health. For controlling the spreading of the virus, we have to prepare for precaution and futuristic calculation for infection spreading. The coronavirus affects the population of the world including Inia. Here, we are the study the virus spreading rate on the Maharashtra state which is part of India. We are predicting the infected people by the SIR model. SIR model is one of the most effective models which can predict the spreading rate of the virus. We have validated the model with the current spreading rate with this SIR model. This study will help to stop the epidemic spreading because it is in the early stage in the Maharashtra region.

## Introduction

A virus[1] is micro/nano meter in size which is reproduced inside the living cell of an organism. Presently coronavirus is created a health emergency to the world population and became a pandemic[2],[3],[4]. Initially, this virus is transferred from the bat animal to the human[5]. Further this virus shows the human to human transmission[6]. COVID 19 virus is spreading to the people by the respiratory droplets and contact mode[7]. Previously there are several mathematical models reported[8]. The SIR model is simple and effective model which can give the prediction of different pandemic situation[9].

Here, we are studying the spreading effects of the COVID19 to the Maharashtra state. There are 28 states in India in which Maharashtra is one of them. This state has a total population of 11.42 crore which is ~8.5 % of the overall population of India. The first case was observed 9 march 2020 in Maharashtra [10] where the couple was returned from Dubai. In India, the first case was observed on 30 January 2020[11]. At 12 march, world health organization (WHO) announce that COVID 19 is outbreak of pandemic where the term used by disease experts when epidemics are growing in multiple countries and continents at the same time[12]. In current situation, after 12 march corona virus cases are rapidly increase in India and there are several cases also observed in the Maharashtra. This study explains the epidemic growth by using SIR Model for this state which helps to control this epidemic.

### Model description

A Susceptible infectious recovered (SIR) model[13],[8] is commonly used for dispersion of the virus as shown in figure 1. This model based upon the population of susceptible *S*, infectious *I*, recovered *Re, N* total Population, *β* Infected ratio, and *γ* Recovery ratio. The set of equation is for this situation.

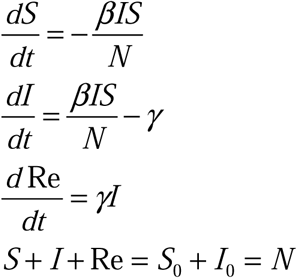

**Figure 1.**
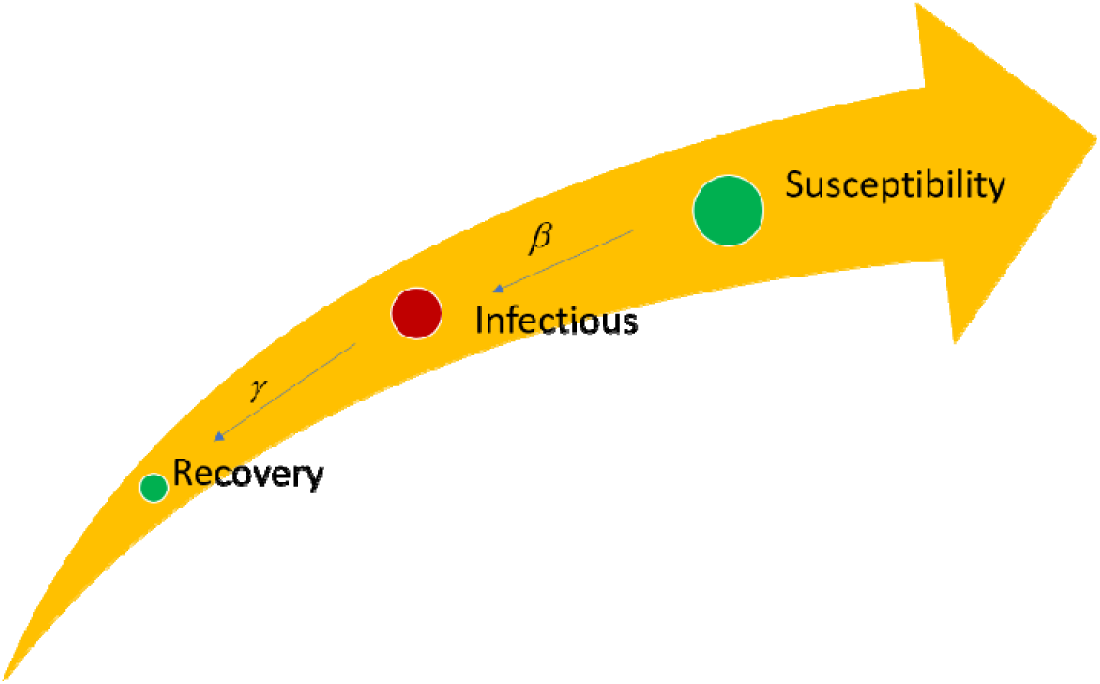
Scheme for SIR model

The ratio of the infected ratio and recovery ration is known as reproduction ratio (*R*).

## Results and Discussions

In the results and discussion section, the present rate of the virus infection is studied. The infected population is also predicted by the SIR model. The reproduction ratio parameter also explained for the spreading rate of the virus.

Presently, India has a total of 4789 in which 1018 cases are from Maharashtra. This state has 21.25 % of the total case of a country. Figure 1 (a) explains the trend of the infected people from day 1 (9 march 2020) to 07 march 2020 by the blackline. The red line shows the death of th people by this virus which is 64 which is 6.28 % of the infected people. This death percentage is a very high compared to the country percentage (2.82 %). The blue line shows the recovery of the people which is 7.76 % of the infected people and the country recovery percentage is 13.56. The individual recovery percentage of the state is low compare to the country percentage. Figure 2 suggests the per week infected people which is increasing continuously. There are large number of people infected in three to four week.

**Figure 2.**
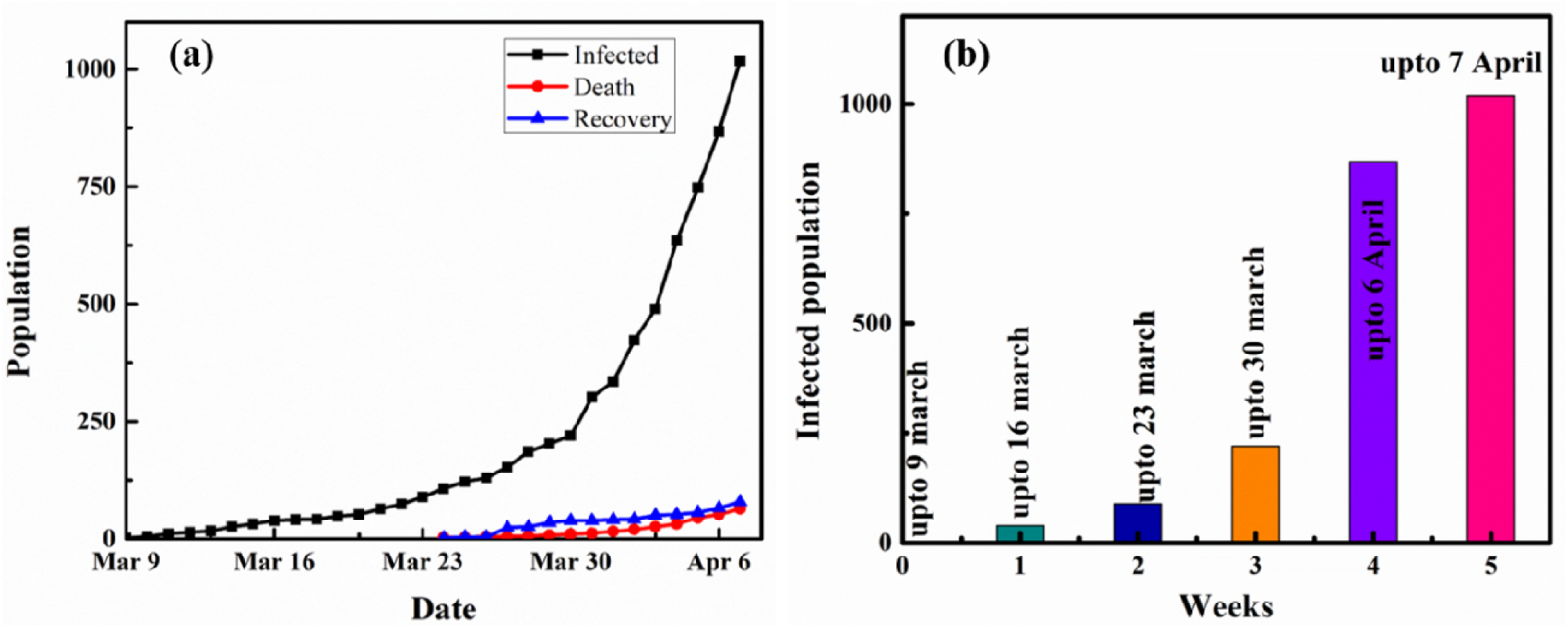
Epidemic situation of Maharashtra **(a)** date wise infected, death and recovery of population **(b)** per week infected people

Figure 3 (a) shows the prediction of the infected population after 7 April 2020. If we make the trendline of the current situation of the infected people. The trend line shows the exponential equation which is

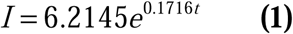

**Figure 3.**
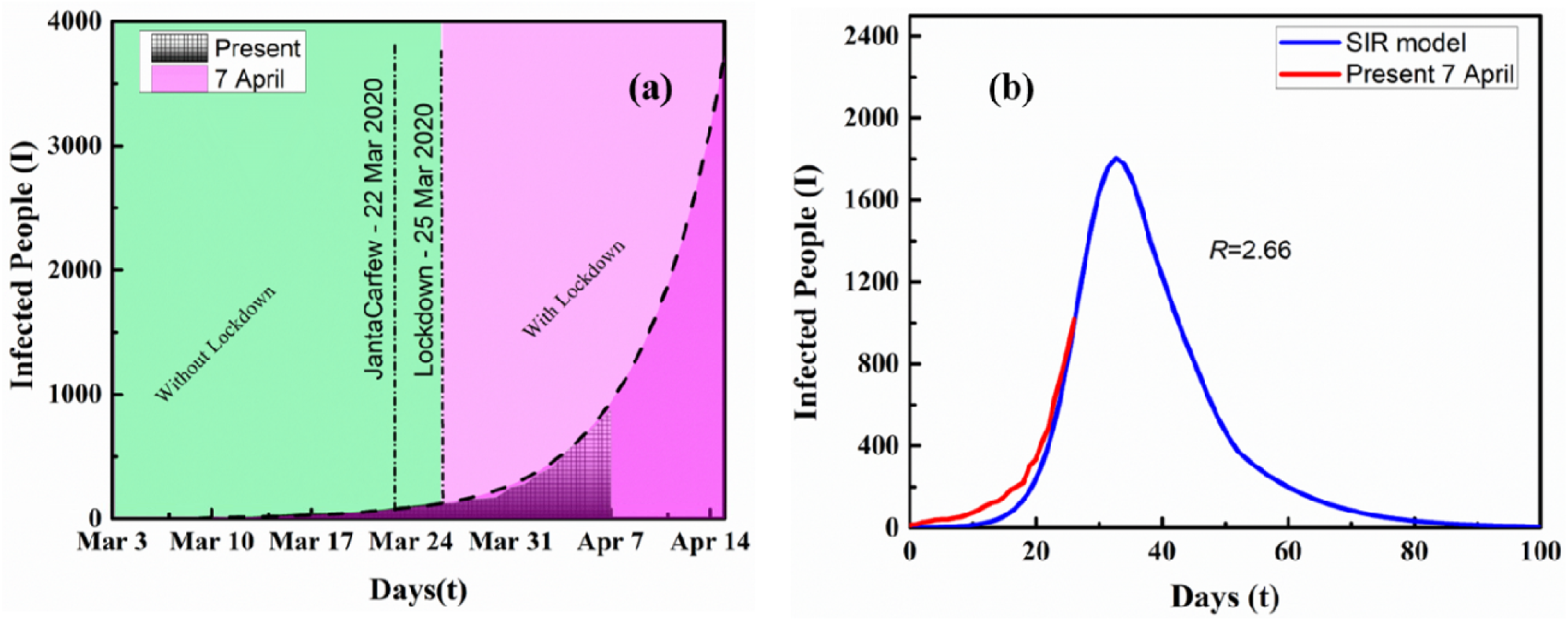
**(a)** Predication of the infected people up to 15 April **(b)** Predication of infected people by SIR model

Where *I* represent the infected people and *t* represents the days.

The undercover area is covered by the black line which follows the equation 1. After that, if we plot the graph up to the 15 April then the infected population will be ~3500. This predication is followed a similar tend when there is no recovery and death of the population.

Figure 3 (b) shows the prediction of infected people by the SIR model which is represented by the black line. The reproduction ratio for this curve is 2.66. Redline is representing the currently infected people which is overlap the black line. This overlapping confirms the infected people will follow this black line curve. After some time, the infected people will less effect by the virus.

SIR model is already explained in the model description section which is applied for the Maharashtra COVID 19 situation. The calculation condition of the SIR model is given into table 1. By using these conditions, there are given three equation solved by the ODE45 in MATLAB. The result is plotted between the susceptible, infected and recover population with different time intervals as shown in figure 4 (a). Blackline represents the susceptible people which is initially not significant changes but after 20 days there are decrease the susceptible people. The infected people are represented by the red line which are initial increased very sharply after reaching a certain peak. After certain period, the infected population are decreases. The blue line shows the recovery or death of the population which is increasing by time. Image 4 (b) shows the reproduction ratio (*R*) effects to the infected people. The reproduction ratio is varied from 1.33 to 5 where infected population was increased in the high reproductive ratio. When susceptible are high than infected people will be more as shown in Figure 4 (c)-(d) and corresponding infected people at different reproduction ratio. During the lockdown, people will make the proper social distancing which will reduce the spreading of infection.

**Figure 4.**
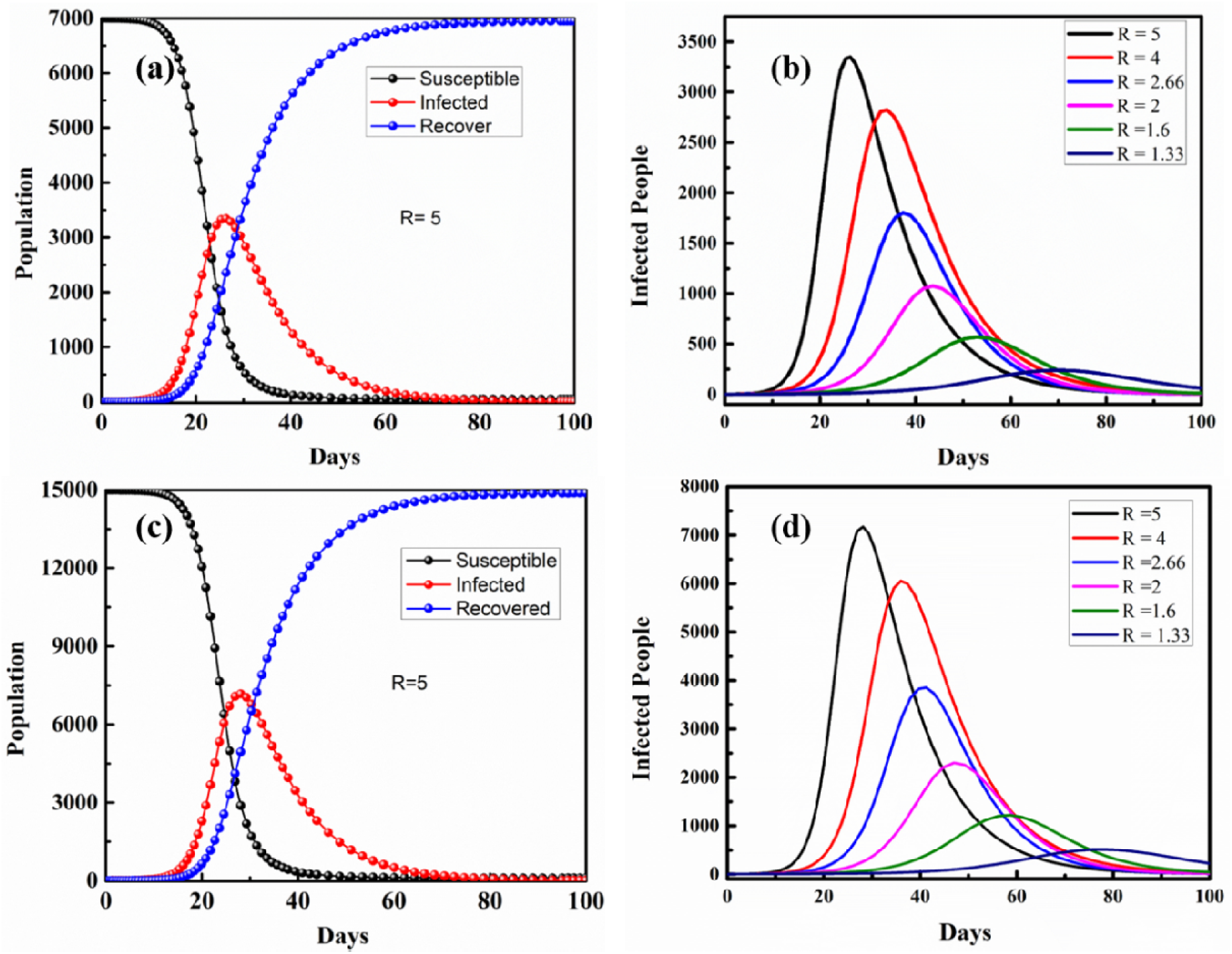
Population verse days plot, **f**or 7000 susceptible **(a)** SIR Model of susceptible, infected and recovered **(b)** Infected people for different reproduction ratio (R) at variable days, for 15000 susceptible **(c)** SIR Model of susceptible, infected and recovered **(d)** Infected people for different reproduction ratio (R) at variable days

**Table 1.** Parameter for SIR model calculation

| Parameter | Values |
| --- | --- |
| $\beta$ | 0.5 |
| $\gamma$ | 0.1 |
| $N$ | 7000 and 15000 |
| $S_o$ | 7000 and 15000 |
| $I_o$ | 1 |
| $R_0$ | 0 |

## Conclusion

The study explains the spreading of COVID 19 in the Maharashtra region which is part of India. In this particular region, the effect of the virus is studied by the SIR model. The model predicted that the maximum infected population will be almost ~ 3000 after 40 days at the peak point. After reaching the peak point the recovery will be more and the virus infection will be high for *R* ~ 2.66. This study is considered only 7000 population are migrating in the state which can be affected in the lockdown condition. If the migrated population will increase which causes the increment of infection and infect the population more. This can control by the lockdown situation only which is presently imposed by the Governments to the people. There should be a global health community that unites to urgently avoid the pandemic issues.

## Data Availability

The data of the number of patients in country is taken from wikipedia. https://en.wikipedia.org/wiki/2020_coronavirus_pandemic_in_Maharashtra https://en.wikipedia.org/wiki/2020_coronavirus_pandemic_in_India https://arogya.maharashtra.gov.in/1175/Novel--Corona-Virus

## Ethical approval

The ethical approval or individual consent was not applicable.

